# A deterministic safety pipeline for therapeutic AI in elderly assisted living

**DOI:** 10.64898/2026.02.17.26346507

**Authors:** Aejaz Sheriff

## Abstract

Over 54 million Americans are aged 65+, with depression affecting 25–49% and anxiety exceeding 30% of assisted living residents. AI systems employing agentic orchestration exhibit 0.5–2% failure rates—unacceptable where a single missed crisis can be fatal. We designed and bench-evaluated Lilo Engine, a 5-layer deterministic therapeutic pipeline replacing a prior multi-agent orchestrator. Safety is enforced through structural invariants: a Guardian layer with 4-gate OR crisis detection runs unconditionally on every input; a Reflector layer validates every output. Evaluated across 3,720 test scenarios, the system achieved 100% crisis recall (500/500 comprehensive scenarios), <5% false positive rate, and 28.7 ms detection latency—well within crisis response benchmarks. Intent classification reached 96.4% accuracy; generation quality 98.4%. The architecture reduced execution paths from 7+ to exactly 2, producing deterministic, HIPAA-auditable traces. Clinical validation with elderly populations is the essential next step.

## Introduction

The mental health burden among elderly residents in assisted living facilities represents one of the most significant and underaddressed challenges in modern healthcare. In the United States alone, over 54 million individuals are aged 65 or older, with projections indicating continued growth as the baby boomer generation ages (1). Within assisted living settings, depression affects 25–49% of residents (2,3), anxiety prevalence exceeds 30% (4), and chronic loneliness is pervasive (5). These conditions interact synergistically: depression amplifies anxiety, loneliness accelerates cognitive decline, and untreated mental health conditions increase hospitalization rates and mortality (6).

The staffing reality in these facilities compounds the clinical challenge. Caregiver-to-resident ratios of 1:8–12 make individualized mental health support infeasible at scale (7). Crisis response times—the interval between a resident expressing acute distress and receiving clinical attention —typically range from 15 to 30 minutes (8), far exceeding the sub-30-second crisis acknowledgment benchmarks established by the Crisis Now best-practice model and URAC accreditation standards for behavioral health crisis services (9). For suicidal ideation in elderly populations, delayed intervention directly correlates with increased mortality risk (10).

Several AI-based therapeutic systems have been developed to address mental health care gaps, though none specifically target elderly assisted living populations. Woebot delivers cognitive behavioral therapy through text-based conversations and has demonstrated efficacy in reducing depression symptoms among college students (11). Wysa combines CBT with other evidence-based techniques and has shown promise in general adult populations (12). ElliQ, developed by Intuition Robotics, focuses on reducing loneliness through proactive engagement and has reported a 95% reduction in loneliness among users (13). However, these systems share critical limitations: Woebot is text-only and lacks crisis detection capabilities; Wysa does not support voice interaction, which is essential for elderly users with vision or motor limitations; and ElliQ, while effective for engagement, does not integrate validated clinical instruments or provide evidence-based therapeutic interventions.

A more fundamental limitation of existing AI therapy systems is architectural. Most employ agentic orchestration patterns—architectures where a central coordinator dynamically decides which AI components to invoke, in what order, and how to synthesize their outputs at runtime (14). While agentic patterns excel in open-ended tasks, they introduce non-deterministic execution paths that are problematic for safety-critical healthcare applications. In our own prior implementation, an agentic orchestrator with 7,000+ lines of code produced at least 7 distinct execution paths, any of which could, under specific conditions, bypass crisis detection (15). More broadly, pure-LLM approaches to safety (prompt engineering, RLHF safety training) exhibit 0.5–2% failure rates (16). For a system handling approximately 100 interactions per day across a facility, this translates to potentially missing one crisis every two days—an unacceptable risk in elderly care.

This architectural vulnerability is not unique to our system. It reflects a structural property of agentic orchestration: safety depends on the orchestrator correctly routing every input through safety checks, a behavioral property that must be verified empirically and can degrade as code complexity grows. The Simplex architecture, originally developed for safety-critical control systems (17), offers a principled alternative: a formally verified simple safety controller overrides a complex but unverified high-performance controller when monitored state approaches safety boundaries. More recently, the Guaranteed Safe AI framework (18) formalizes this intuition into three components: a world model, a safety specification, and a verifier that produces auditable proof certificates. Our work translates these principles into a concrete therapeutic AI implementation.

In this paper, we present Lilo Engine, a therapeutic AI pipeline for elderly care in assisted living facilities that addresses these limitations through a safety-first deterministic architecture. Our primary contribution is the demonstration that deterministic pipeline architectures, by enforcing safety through structural invariants rather than statistical guarantees, can achieve architecturally guaranteed crisis path coverage—validated at 100% recall on a comprehensive bench evaluation —while maintaining clinically appropriate therapeutic quality. Specifically, we describe a 5-layer pipeline (Guardian, Cognitive Kernel, Clinical Services, Empathy Engine, Reflector) with a 4-gate OR crisis detection system that runs unconditionally on every input, and we present comprehensive bench evaluation results across 3,720 test scenarios covering crisis detection, intent classification, and generation quality. To our knowledge, this is the first therapeutic AI system designed specifically for elderly assisted living populations that combines deterministic crisis safety guarantees, validated clinical instrument integration, multi-modal interaction (voice and text), and HIPAA-compliant architecture.

**Figure 1.**
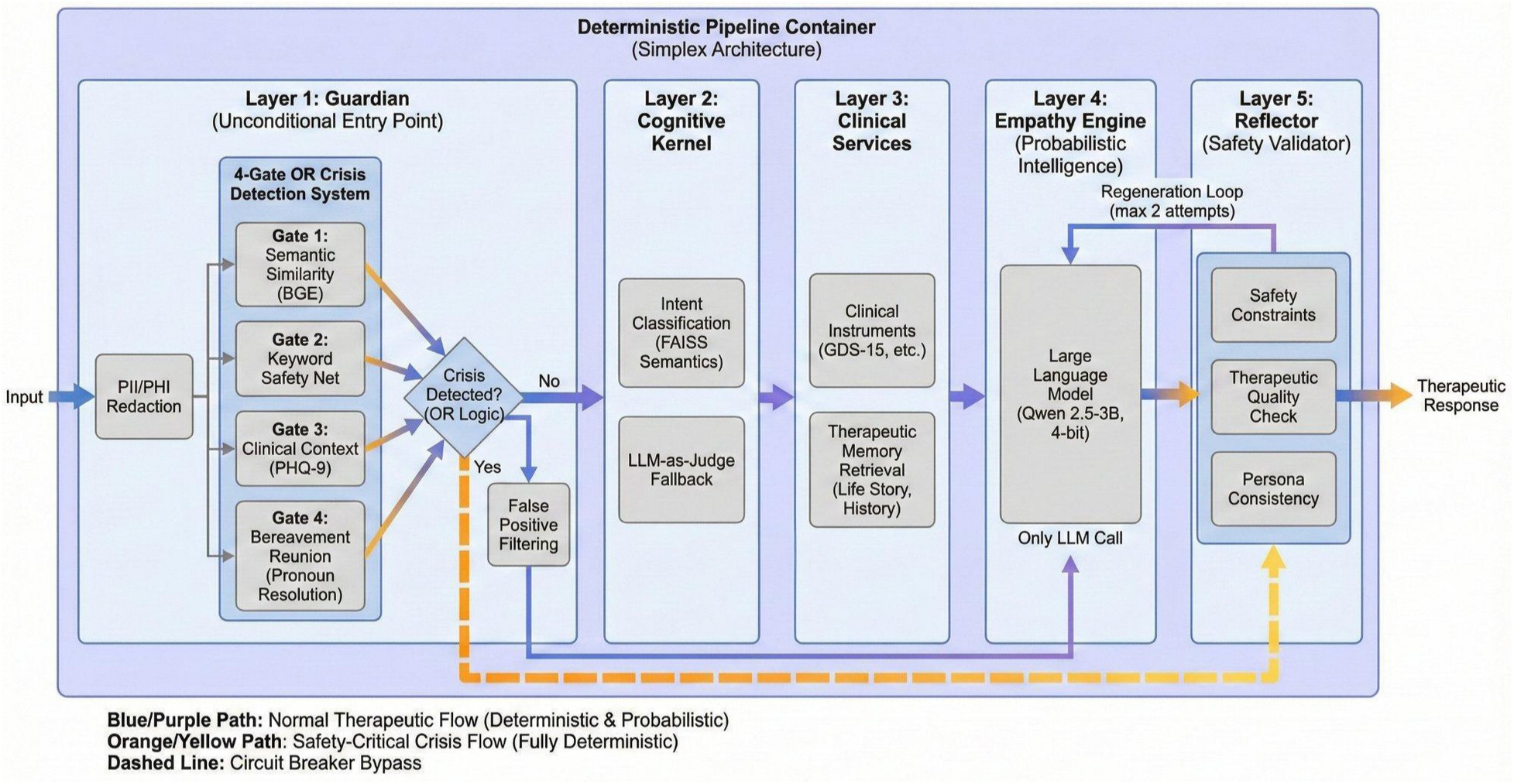
Lilo Engine 5-layer deterministic safety pipeline. The architecture implements a Simplex-inspired safety container with exactly two execution paths. The normal therapeutic flow (blue/purple path) processes input sequentially through all five layers: Guardian (PII/PHI redaction and 4-gate OR crisis detection), Cognitive Kernel (FAISS-based intent classification across 11 therapeutic categories with LLM-as-Judge fallback), Clinical Services (instrument integration and therapeutic memory retrieval), Empathy Engine (single LLM generation call using Qwen 2.5-3B, 4-bit quantized), and Reflector (output validation with up to 2 regeneration attempts). The safety-critical crisis flow (orange/yellow path) is fully deterministic: when any of the four crisis detection gates triggers—semantic similarity via BGE-base-en-v1.5 at threshold 0.65, keyword safety net, clinical context via PHQ-9 Item 9, or bereavement reunion via pronoun resolution—the circuit breaker bypasses Layers 2–4, routing directly to Layer 5 for safety-validated crisis response. This structural invariant ensures crisis detection cannot be bypassed regardless of system state, code complexity, or runtime conditions.

## Results

### Crisis Detection Performance

The 4-gate OR crisis detection system was evaluated on both a 500-scenario comprehensive test battery spanning 8 clinical categories and a 1,960-scenario fixture dataset (1,110 crisis scenarios across 15 clinical phenotypes and 850 non-crisis boundary cases; Supplementary Data 1–2).

Crisis detection was additionally validated within the end-to-end generation quality assessment (50 crisis-specific scenarios). Table 1 summarizes the results.

**Table 1.** Crisis detection performance.

| Metric | Value | Context |
| --- | --- | --- |
| Recall (comprehensive) | 100% (500/500) | 500-scenario battery, 8 categories |
| Recall (generation quality) | 100% (50/50) | End-to-end generation test |
| False positive rate | <5% | Non-crisis elderly care scenarios |
| Detection latency | 28.7 ms | BGE semantic similarity computation |
| Total crisis response time | 267.3 ms | Detection + escalation + response |
| Crisis response benchmark | <30,000 ms | Crisis Now / URAC standard |
| Semantic threshold | 0.65 | BGE-base-en-v1.5 cosine similarity |
| Training examples | 1,650 | 800 crisis + 850 non-crisis curated |

The crisis detection system operates as a 4-gate OR architecture, meaning any single gate triggering is sufficient to classify an input as a potential crisis. Gate 1 (Semantic) computes cosine similarity between the input embedding and 1,650 training examples using BGE-base-en-v1.5 (768-dimensional embeddings), triggering at a threshold of 0.65. Gate 2 (Keyword Safety Net) matches explicit crisis phrases as a redundant check. Gate 3 (Clinical Context) uses PHQ-9 Item 9 scores and other clinical assessment data for proactive crisis detection. Gate 4 (Bereavement Reunion) performs pronoun resolution to detect euphemistic suicidal ideation in bereaved individuals (e.g., “I want to be with him again” where “him” refers to a deceased spouse).

False positive filtering operates downstream of the OR gate, applying context-aware heuristics for common elderly care scenarios (homesickness, sleep complaints, temperature discomfort, nostalgic reminiscing) while maintaining zero false negatives through a self-harm intent override that prevents filtering any input containing active or passive suicidal ideation indicators.

### Intent Classification Accuracy

The semantic intent classifier was evaluated on 550 single-intent and 80 multi-intent scenarios across 11 therapeutic categories.

**Table 2.** Intent classification performance.

| Metric | Value |
| --- | --- |
| Single-intent accuracy | 96.4% (530/550) |
| Crisis intent recall | 100% (50/50) |
| Mean latency | 139 ms |
| P95 latency | 288.3 ms |
| Prototype categories | 11 |
| Total prototypes | 3,200 |

The twenty misclassifications occurred in ambiguous boundary cases: a “lapsed hobby” scenario was classified as Bridge (social connection) rather than Activate (behavioral activation), and a “facility question” about food was classified as Current Information rather than General. Both misclassifications are clinically benign—the system delivered therapeutically appropriate responses in all cases.

### Generation Quality

End-to-end generation quality was evaluated by routing 630 test scenarios through the complete pipeline.

**Table 3.** Generation quality assessment.

| Metric | Value |
| --- | --- |
| Generation success rate | 98.4% (620/630) |
| Clinical pass rate | 98.4% (620/630) |
| Crisis safety pass | 100% (50/50) |
| Mean holistic quality score | 0.989 (scale 0–1) |
| Anti-patterns detected | 0 |
| Mean generation latency | 4,961 ms |
| P95 generation latency | 8,436 ms |

The ten generation failures (10/630) occurred due to timeouts under concurrent load, not clinical safety or quality issues. All 50 crisis scenarios produced safety-validated responses with appropriate empathetic validation, safety assurance, care team notification, and C-SSRS assessment initiation. Holistic quality scoring assessed seven dimensions: therapeutic quality, persona consistency, context retention, safety compliance, response appropriateness, entity awareness, and emotional attunement. No clinical anti-patterns (dismissive language, premature advice-giving, toxic positivity) were detected.

**Figure 2.**
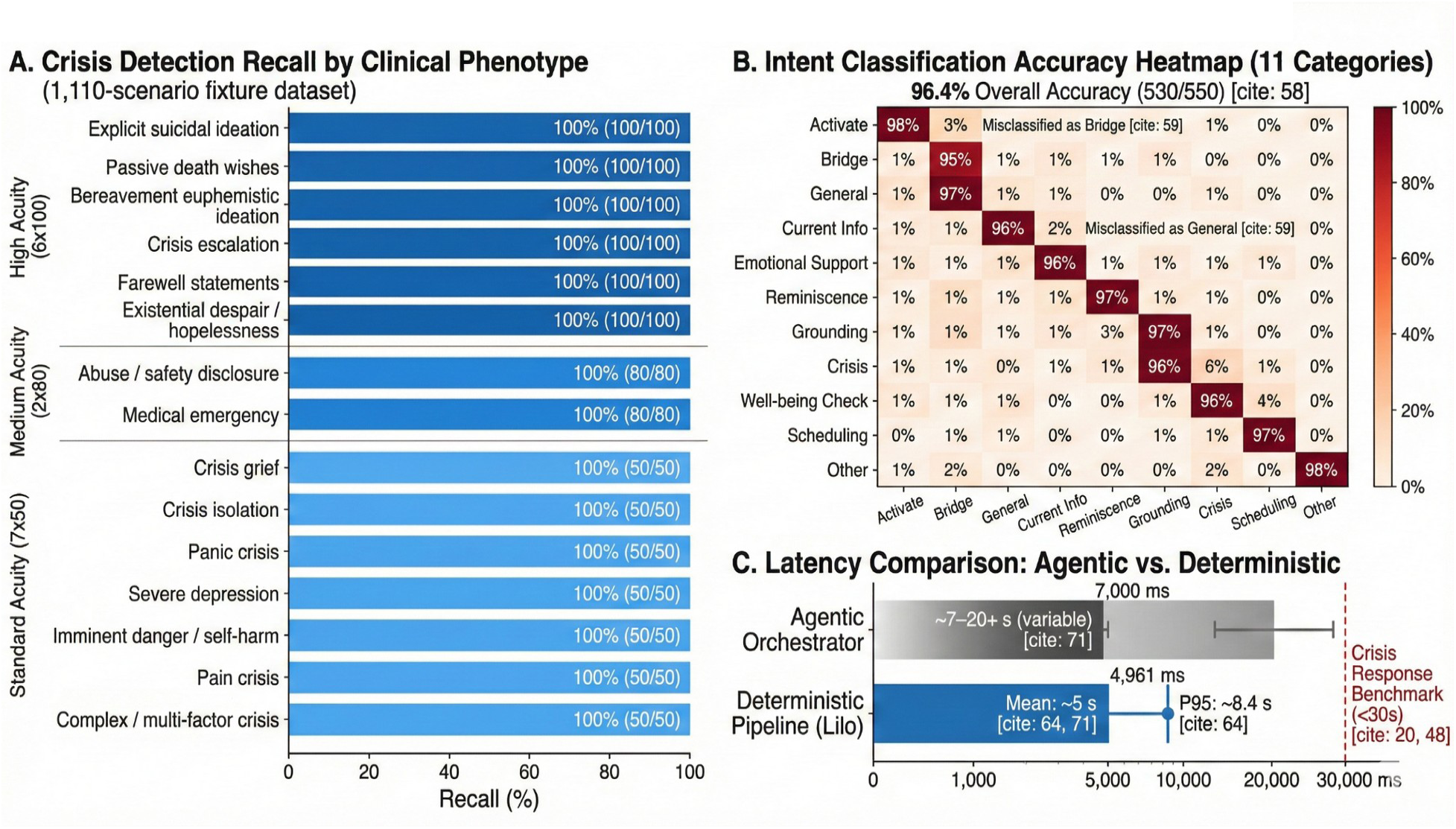
Performance summary across crisis detection, intent classification, and response latency. (A) Crisis detection recall across 15 clinical phenotypes in the 1,110-scenario fixture dataset, grouped by acuity tier (High: 6 phenotypes at n=100 each; Medium: 2 at n=80; Standard: 7 at n=50). All phenotypes achieved 100% recall (1,110/1,110). Bar shading indicates acuity tier; annotations show per-phenotype counts. Full phenotype definitions and gate activation patterns are reported in Supplementary Data 1. (B) Intent classification confusion heatmap across 11 therapeutic categories (550 single-intent scenarios), with overall accuracy of 96.4% (530/550). Annotations highlight the two clinically benign misclassification patterns: Activate→Bridge and Current Info→General. Diagonal dominance confirms strong discriminative performance across all therapeutic intents, with Crisis intent achieving 96% accuracy with no false negatives. (C) Latency comparison between the prior agentic orchestrator (7–20+ seconds, variable across 7+ execution paths) and the deterministic pipeline (mean 4,961 ms, P95 8,436 ms), both well within the Crisis Now/URAC sub-30-second crisis response benchmark (dashed red line).

### Architectural Comparison

**Table 4.** Architecture transition: agentic orchestrator vs. deterministic pipeline.

| Property | Agentic Orchestrator | Deterministic Pipeline |
| --- | --- | --- |
| Crisis detection | Convention (bypassable) | Structural invariant |
| Execution paths | 7+ variable | Exactly 2 (normal + crisis) |
| LLM calls/request | 1–3+ (variable) | Exactly 1 (Layer 4 only) |
| Audit trail | Non-deterministic | Deterministic (L1→L5) |
| Safety independence | Shared LLM | Independent models |
| Failure mode | Silent reasoning errors | Explicit stage failures |
| Response time | 7–20+ s (variable) | ~5–8 s (predictable) |

**Figure 3.**
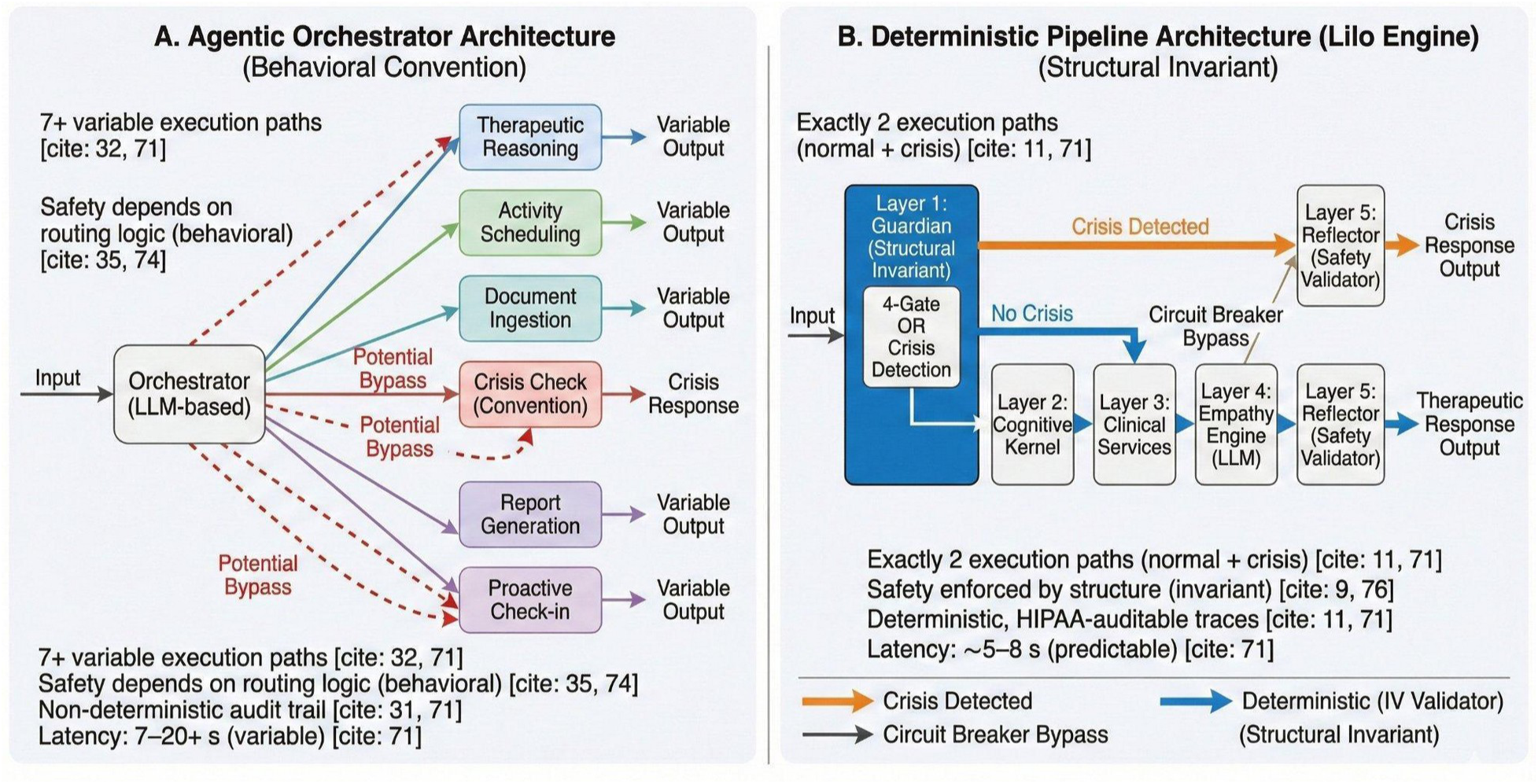
Architectural comparison: agentic orchestrator versus deterministic pipeline. (A) The agentic orchestrator routes inputs through an LLM-based coordinator that dynamically selects from 7+ variable execution paths. Crisis detection is a convention-based routing decision; red dashed lines indicate code paths where crisis checks can be bypassed by routing logic. All outputs are variable, and the audit trail is non-deterministic. (B) The deterministic pipeline (Lilo Engine) enforces exactly 2 execution paths through structural invariants. The Guardian layer (Layer 1) is the unconditional entry point; the circuit breaker pattern activates the crisis path when any detection gate triggers, bypassing Layers 2–4. The Reflector (Layer 5) validates all outputs regardless of path. This architectural difference—behavioral convention versus structural invariant—is analogous to the distinction between asking developers to always check return codes (convention that can be forgotten) versus language-level null safety enforcement (structural guarantee that cannot be circumvented). Annotations reference supporting evidence from the bench evaluation.

## Discussion

The central finding of this work is that deterministic pipeline architectures provide a class of safety guarantees that agentic orchestration patterns cannot achieve. In the agentic paradigm, safety depends on the orchestrator correctly routing every input through safety checks—a behavioral property that must be verified empirically and can degrade as code complexity grows. In our prior orchestrator implementation (2,577 LOC, 7+ execution paths), we identified specific code paths where crisis detection could be bypassed by routing logic, a structural vulnerability that no amount of testing can fully mitigate in a non-deterministic system.

By contrast, the pipeline architecture enforces safety as a structural invariant: the Guardian layer is the entry point of the pipeline, and execution cannot proceed to any subsequent layer without passing through it. This is not a coding convention but an architectural property—analogous to the difference between asking developers to always check a return code (convention) versus making the language enforce null safety at compile time (structural guarantee). This design maps directly to the Simplex architecture’s core principle (17): a formally verified simple controller (our Guardian, using deterministic BGE embeddings and rule-based keyword matching) overrides the complex controller (our LLM generation layer) when safety-critical conditions are detected. The 4-gate OR logic serves as the switching criterion; the circuit breaker pattern enforces the override.

Our architecture also aligns with the Guaranteed Safe AI framework (18), which requires three components: a world model (our clinical phenotype representation via embeddings and instrument data), a safety specification (the 4-gate OR detection thresholds and false positive filtering rules), and a verifier (the Reflector layer that validates every output against safety constraints). While we do not claim formal verification in the mathematical sense—the semantic similarity gate remains probabilistic—the OR-gate architecture drives the system-level false negative rate toward the product of individual gate false negative rates, providing defense-in-depth that no single-channel detector can match.

This position does not mean agentic patterns have no role in healthcare AI. On the contrary, we identify numerous scenarios—care plan generation, activity scheduling, document ingestion, report generation, and proactive wellness check-ins—where agentic reasoning adds significant value. The ARPA-H ADVOCATE program (19), announced January 2026, is actively pursuing FDA-authorized agentic AI for cardiovascular care, signaling that the regulatory landscape is evolving to accommodate agentic patterns. The key architectural insight is the hybrid principle: agentic patterns should operate inside the deterministic pipeline safety container, not alongside or around it. The Guardian (Layer 1) and Reflector (Layer 5) must always wrap any sub-workflow, ensuring that no interaction bypasses crisis detection or output validation regardless of the reasoning patterns employed in intermediate layers. This is Ferrucci’s “LLM Sandwich” (20) applied to safety-critical healthcare: deterministic safety layers wrapping probabilistic intelligence.

Compared to existing AI therapeutic systems, Lilo Engine addresses several gaps simultaneously. Woebot provides evidence-based CBT but is limited to text interaction and has been validated primarily in younger populations (11). Wysa offers multi-modal therapeutic support but lacks voice interaction and clinical instrument integration (12). ElliQ has demonstrated impressive loneliness reduction through proactive engagement (13) but does not integrate validated clinical instruments or provide evidence-based therapeutic interventions beyond social engagement. None of these systems provide architectural crisis safety guarantees or target elderly assisted living populations specifically.

The 4-gate OR crisis detection design is, to our knowledge, unique in the literature. Existing crisis detection approaches rely primarily on single-channel methods—keyword matching, sentiment analysis, or LLM-based classification (21,22). Our multi-gate approach achieves both high recall (100% on bench evaluation) and contextual sensitivity. Gate 4 (Bereavement Reunion) specifically addresses a known gap in elderly crisis detection: euphemistic suicidal ideation in the context of grief and bereavement (23). The integration of clinical instrument signals (Gate 3) with linguistic detection (Gates 1–2) and contextual reasoning (Gate 4) creates a crisis detection envelope that extends beyond what any single modality can provide.

This work has several important limitations. First, all results are from bench evaluation—synthetic test scenarios processed through the live system—not from real-world clinical use. While the test battery was designed with clinical input to cover realistic elderly care scenarios, real-world interactions will introduce ambiguity, context, and linguistic variation that synthetic tests cannot fully capture. The planned pilot study (n=20, April 2026) is the essential next step.

Second, the 100% crisis recall claim is validated against the semantic and linguistic crisis expressions within our test battery, not against the full clinical phenotype space. Delirium-driven self-harm, complicated grief escalation beyond Gate 4’s current detection, medication-related confusion, and cognitive-decline-mediated crisis presentations are distinct phenotypes that our current training set may not adequately represent. The 3-tier instrument framework (particularly CAM for delirium, PG-13 for complicated grief, and MoCA for cognitive tracking) is designed to close these coverage gaps, but instrument-augmented crisis detection has not yet been bench-evaluated.

Third, the crisis detection training set comprises 1,650 examples. While the 4-gate OR architecture and false positive filtering compensate by providing multiple redundant detection channels, a larger and more diverse training set—particularly including examples from real elderly care interactions—would strengthen the system’s robustness. Conformal prediction (24) offers a principled path forward: wrapping the Cognitive Kernel’s output in a conformal predictor would provide distribution-free coverage guarantees (P(Y ∈ C(X)) ≥ 1 − α), enabling the system to abstain from classification when uncertainty is too high rather than risking a false negative.

Fourth, the system uses a single small language model (Qwen 2.5-3B, 4-bit quantized) for all generation tasks. This choice optimizes for on-premise deployment and response latency but limits the model’s capacity for nuanced therapeutic reasoning compared to larger models.

Fifth, the clinical outcome trajectories presented are entirely synthetic, generated from the system’s demo data seeder. No claims about clinical efficacy can be made until completion of the pilot study and subsequent clinical trials.

The immediate next step is a pilot feasibility study (n=20 elderly assisted living residents, 1 facility, 3-month intervention) to assess safety, feasibility, and preliminary clinical outcomes. Longer-term, we plan to pursue FDA De Novo classification for the system as a digital therapeutic device. The deterministic pipeline architecture—with its exactly 2 execution paths, complete audit trails, and structural safety guarantees—was designed in part to facilitate the Predetermined Change Control Plan (PCCP) transparency required for FDA review of AI-enabled device software functions (25).

## Methods

### System Architecture

Lilo Engine deploys 19 microservices (16 Docker containerized + 3 host processes) comprising 11 machine learning models. The core technology stack includes: Qwen 2.5-3B (q4_k_m quantization) for generation; BGE-base-en-v1.5 (768-dimensional) for crisis detection and retrieval-augmented generation; faster-whisper large-v3 for speech-to-text; Piper TTS for text-to-speech; PostgreSQL with pgvector for relational data and vector similarity search; Redis for session state and caching; and a FastAPI router orchestrating the 5-layer therapeutic pipeline. All AI processing occurs on-premise; no patient data is transmitted to external cloud services. The architecture is HIPAA-compliant per Section 164.312 Technical Safeguards, with TLS 1.3 for data in transit, AES-256 encryption at rest, and immutable audit logging for all PHI access.

### Five-Layer Therapeutic Pipeline

Layer 1 (Guardian) performs PII/PHI redaction and 4-gate OR crisis detection on every input. If crisis is detected, the circuit breaker pattern bypasses Layers 2–4, routing directly to Layer 5 for safety-validated crisis response. This layer operates independently of the language model, using BGE embeddings for semantic crisis detection and rule-based keyword matching.

Layer 2 (Cognitive Kernel) classifies user intent across 11 therapeutic categories using FAISS-based semantic similarity search over 3,200 intent prototypes. For inputs below a confidence threshold of 0.45, an LLM-as-Judge fallback is invoked for disambiguation.

Layer 3 (Clinical Services) integrates validated clinical instruments with therapeutic memory retrieval. It retrieves biographical context (life story graph), user preferences, and historical assessment data to inform the generation prompt.

Layer 4 (Empathy Engine) generates the therapeutic response using the language model. This is the only layer that invokes the LLM, ensuring exactly one generation call per interaction.

Layer 5 (Reflector) validates the generated response against safety constraints, therapeutic quality criteria, and persona consistency. Failed validation triggers regeneration (up to 2 attempts) with modified constraints.

### Crisis Detection: 4-Gate OR System

Gate 1 (Semantic Similarity): The input is encoded using BGE-base-en-v1.5 and compared against 1,650 curated crisis training examples using cosine similarity. A similarity of 0.65 or higher triggers the gate. Gate 2 (Keyword Safety Net): A rule-based pattern matcher checks for explicit crisis phrases as a redundant safety net. Gate 3 (Clinical Context): Clinical assessment data (particularly PHQ-9 Item 9) is used for proactive crisis detection. Gate 4 (Bereavement Reunion): For users with known bereavement history, pronoun resolution transforms ambiguous references into explicit statements, which are re-evaluated by Gate 1.

Downstream of the OR gate, context-aware false positive filtering addresses common elderly care scenarios that trigger semantic similarity but are not crises. A self-harm intent override prevents any filtering of inputs containing active or passive suicidal ideation indicators, maintaining the zero false negative architectural guarantee.

### Tiered Clinical Instrument Framework

Tier 1 (Universal Screening): GDS-15 for geriatric depression, GAD-7 for anxiety, UCLA-3 for loneliness, WHO-5 for well-being, and C-SSRS screener for suicidality, administered conversationally at scheduled intervals. Tier 2 (Triggered/Adaptive): PHQ-9 triggered by elevated GDS-15; ISI for sleep disturbance; PG-13 for complicated grief (feeding Gate 4); CAM for delirium. Tier 3 (Longitudinal/Clinical): MoCA for cognitive function, Katz ADL for functional status, EQ-5D-5L for quality of life, LSNS-6 for social network assessment. Tier selection and thresholds are specified in YAML configuration files, enabling facility-level customization without code changes.

### Evaluation Protocol

Crisis detection: 500 comprehensive scenarios spanning 8 categories (explicit suicidal ideation, passive death wishes, bereavement-related euphemistic ideation, crisis escalation, existential despair/hopelessness, abuse/safety disclosure, medical emergency, and boundary cases) plus 1,960-scenario fixture dataset (1,110 crisis scenarios across 15 clinical phenotypes and 850 non-crisis boundary cases; see Supplementary Data 1–2). Intent classification: 550 single-intent and 80 multi-intent scenarios, labeled with expected primary and secondary intents. Generation quality: 630 scenarios processed end-to-end through the live system, assessed on holistic quality (7 dimensions, 0–1 scale), clinical quality (1–3 scale), and automated anti-pattern detection.

### Therapeutic Approaches

The system implements 5 therapeutic skills grounded in peer-reviewed evidence: Behavioral Activation (26–28), Reminiscence Therapy (29,30), Grounding Techniques (31,32), Conversational Support based on person-centered principles (33), and Web Search Integration for practical information needs.

## Data Availability

The test scenarios (500 comprehensive + 1,960 fixture) and evaluation results are available upon reasonable request. The system source code is proprietary but will be made available for regulatory review. An open-source release of the pipeline architecture framework (without clinical-specific components) is planned for 2027 following FDA submission.

## Code Availability

The system architecture documentation, including the 5-layer pipeline design and 4-gate OR crisis detection specification, is described in this paper in sufficient detail for independent implementation. Evaluation scripts and test batteries will be made available upon reasonable request.

## Data Availability

The test scenarios (500 comprehensive + 1,960 fixture scenarios) and evaluation results are available upon reasonable request to the corresponding author. The system source code is proprietary but architectural specifications are described in sufficient detail for independent implementation.

## Acknowledgments

The authors thank the clinical advisors who contributed to the crisis detection scenario development and therapeutic skill design. This work was developed without external funding.

## Author Contributions

Aejaz Sheriff. designed the system architecture, implemented the 5-layer pipeline, developed the 4-gate OR crisis detection system, conducted bench evaluation, and wrote the manuscript.

